# A single center retrospective study to examine the effect of concomitant metformin treatment on cisplatin induced nephrotoxicity in adult HNSCC patients between 2015-2021

**DOI:** 10.1101/2024.06.14.24308941

**Authors:** Dawud Ellayan

**Author notes:** This research did not receive any specific grant from funding agencies in the public, commercial, or not-for-profit sectors.

## Abstract

**Purpose:** Examine the effect of AMPK activation in addition to OCT2 competitive blockage through metformin concomitant treatment on the incidence rate of nephrotoxicity in adult head and neck cancer patients treated with cisplatin-based chemoradiation.

**Methods:** A single center retrospective three to one controlled study in HNSCC patients treated at a single academic health center between January 1st 2015 to December 31st 2021. Patients treated with cisplatin based chemoradiation regimen at a dose of either 40 mg/m2 weekly, or 100 mg/m2 every 3 weeks for a total of 7 weeks were identified and were divided into two cohorts; Cohort A with patients who received concomitant metformin therapy, where concomitant is defined as taken prior to the time of cisplatin start and continued during treatment. And cohort B with a control group of patients who did not receive metformin during cisplatin treatment.

**Results:** 18 patients were enrolled retrospectively in cohort A and 54 in cohort B. Our data shows a lower incidence of nephrotoxicity than reported in historical controls. However, no statistically significant differences were identified in direct comparison between the two cohorts.

**Conclusion:** Our data reaffirms the higher risk of nephrotoxicity for patients on Q3weeks regimen compared to weekly regimen, however, we were unable to show a statistically significant effect in direct comparison between the cohorts due to sample size limitation.

## Introduction

Cisplatin is a widely used chemotherapeutic agent for the treatment of different solid tumors including in the first and second-line settings in head and neck squamous cell carcinoma (HNSCC). However, treatment is often complicated by drug induced toxicities such as nephrotoxicity which is seen in around third of patients in a dose dependent manner ^1–3^. This contributes to treatment delay and discontinuation in approximately 70% of patients treated with weekly cisplatin and it results in dose reduction in a quarter of patients treated with every three weeks regimen^4–6^. Cisplatin induced nephrotoxicity is due to uptake of cisplatin into renal epithelial cells which result in injury to mitochondrial and nuclear DNA leading to activation of cell death pathways which is a companied by a strong inflammatory response and injury ^3^. Hence, nephrotoxicity prevention can be theoretically accomplished by either inhibiting uptake of cisplatin into renal epithelial cells or through inhibition of steps leading to cell death or a combination of the two.

First, uptake of Cisplatin into renal epithelial cells is governed by Organic cation transporter 2 (OCT2) and multidrug and toxin extrusion protein 1 (MATE1) ^7–9^. While OCT2 is involved in cisplatin uptake by renal epithelial cell, MATE1 is involved in excretion of cisplatin outside of the cell ^10–12^. As such, it is important for any potential drug examined for cisplatin nephrotoxicity prevention to block OCT2 without affecting MATE1 or in an ideal situation block OCT2 transported while inducing expression of MATE1. Evidently, several animal studies have shown that blocking OCT2 reduces cisplatin cellular uptake and consequent toxicity^7,8,13–16^. Furthermore, Lower risk of nephrotoxicity is seen in patients with OCT2 gene polymorphism^17^. Thus, targeting cisplatin transport into or out of the renal epithelial cells represents a valid approach for prevention of cisplatin induced nephrotoxicity.

Second, prevention of renal cell death after uptake of cisplatin by preservation of mitochondrial function though induction of MP-activated protein kinase (AMPK) complex could protect against cisplatin induced renal toxicity. AMPK is an important regulator of Mitochondrial health in response to stressors by regulating energy production and consumption inside the cell^18^. Furthermore, AMPK plays an important role as regulator of active transport process in the kidney^19^. As such, regulation of AMPK signaling might confer protection against cisplatin induced renal toxicity^20^. Indeed, various pre-clinical studies have shown the protective effect of AMPK induction against cisplatin nephrotoxicity^21–25^. Consequently, drugs that are known to induce AMPK signaling are expected to play a protective role against cisplatin induced nephrotoxicity.

Our study aimed to examine the effect of AMPK activation in addition to OCT2 competitive blockage on the incidence rate of nephrotoxicity in adult head and neck cancer patients treated with cisplatin-based chemotherapies. This is accomplished by examining the effect of concomitant treatment with metformin on nephrotoxicity rate in newly diagnosed HNSCC patients. Metformin was selected as it is an extensively used type 2 diabetes medication, that can theoretically reduce cisplatin nephrotoxicity through several pathways^26^. Primarily, it can exert a protective role against cisplatin induced nephrotoxicity through stimulating AMPK activation in the tubular cells^27,28^. Moreover, metformin is known to be an OCT2 substrate ^29^ as such it may also prevent cisplatin uptake by renal epithelial cells through competitive inhibition. Thus, metformin may prevent cisplatin induced renal toxicity. On another note, since metformin is mainly used to treat type 2 diabetes, concerns may be warranted over the potential confounding effect of diabetes on the protective effect of metformin. However, studies have shown no correlation between diabetes and cisplatin nephrotoxicity^30,31^. Hence, the potential for diabetes to confound our results is minimal. A retrospective study design is used due to feasibility concerns with a prospective study in the desired population.

## Methods

### Study Design and Setting

A single center retrospective three to one controlled study to examine the effect of concomitant treatment with metformin on cisplatin induced nephrotoxicity in adult HNSCC patients. Patients treated with cisplatin based chemoradiation regimen at a dose of either 40 mg/m^2^ weekly, or 100 mg/m^2^ every 3 weeks for a total of 7 weeks between January 1^st^ 2015 to December 31^st^ 2021 were identified through the Data Direct system^32^. Patients were divided into two cohorts; Cohort A with patients who received concomitant metformin therapy, where concomitant is defined as taken prior to the time of cisplatin start and continued during treatment. And cohort B with a control group of patients who did not receive metformin during cisplatin treatment. Subsequently patients’ data was obtained through review of the electronic medical record. The retrieved data included cisplatin regimen and duration, Magnesium (Mg), Serum creatinine (Scr) and blood urea nitrogen (BUN) levels at baseline and at the end of treatment, incidence of reported nephrotoxicity, smoking status, presence of hypertension or cirrhosis, use of amifostine, mannitol or magnesium sulfate, and hydration to prevent nephrotoxicity.

## Ethical Considerations

This study was submitted to Michigan Medicine IRB and was granted a waiver (HUM00219934).

### Statistical analysis

Statistical comparisons between the treatment cohort (A) and the control cohort (B) were done using the Mann–Whitney U test and Fisher’s exact test for continuous and categorical variables, respectively. Furthermore, multivariate logistic regression analysis was used to identify the impact of co-administration of metformin on Scr levels when given concomitantly with cisplatin. The logistic regression was adjusted for the following, factors: age, sex, smoking, hypertension, cirrhosis, as they could a potentially have a confounding role^3,6^. Furthermore, Wilson’s formula was used to calculate confidence intervals for proportions. Statistical tests were completed and verified using R software, Datatab software, and Excel.

## Results

During the study period January 1st 2015 to December 31st 2021, 72 HNSCC patients with stage I-IVb disease were identified to meet the study entry requirements, of which 18 patients were enrolled in cohort A and 54 in cohort B. Demographic and clinical data for the two cohorts are presented in table 1 in appendix A. No significant differences between the two cohorts’ characteristics were identified. On another note, all patients received a regimen comprised of hydration, magnesium sulfate and mannitol with their cisplatin doses, but none of the patients received amifostine. As for the chemotherapy regimen used; 12 patients in cohort A and 24 patients in cohort B received chemoradiation with cisplatin 100 mg/m^2^ every 3 weeks (Q3Weeks) for 7 weeks, while the remaining patients from each group receiving chemoradiation with cisplatin 40 mg/m^2^ weekly for 7 weeks.

Patients in cohort A were 40% less likely to discontinue treatment early for any reason (95% CI 0.2700 to 1.3332 P = 0.209). Furthermore, nephrotoxicity risk for patients treated with the Q3week regimen was 60% lower in patients in cohort A (8%) than in cohort B (21%). Moreover, the overall change in serum creatinine (Scr) from baseline to end of treatment was 0.04 mg/dL (4.11%) in cohort A versus 0.14 mg/dL (16.16%) in cohort B (U = 403. Z-score=1.07288. P=0.28462). However, none of these results are statistically significant. On the other hand, there is a statistically significant increase in Scr from baseline in all patients treated with Q3weeks cisplatin regimen vs the weekly regimen regardless of metformin status (P .007 95% CI 0.08-.048). Lastly, no notable changes in Mg levels nor BUN levels were seen between the two cohorts.

## Discussion

In this study, we explored the effect of concomitant treatment with metformin on nephrotoxicity rate in patients with HNSCC treated with chemoradiation. Our study is a three to one controlled retrospective design which shows a trend toward benefit with a reduction in all cause treatment discontinuation and lower incidence of nephrotoxicity in patients treated with cisplatin 100 mg/m^2^ Q3weeks. Furthermore, the change in serum creatinine from treatment start to end was lower in the metformin group. Which is promising even though it was not statistically significant when compared to cohort B patients, since we showed a rate of nephrotoxicity of 8% (95% CI 1.5%-35%) in metformin group vs 17% (CI 8%-32%) in control group in patients treated with the Q21days regimen which are significant reductions when compared to a historical reported rate of renal toxicity of 40% of HNSCC patients treated with same regimen of chemoradiation^33^.

On the other hand, comparison against historical control is not appropriate as many differences that might exist, it is worth noting that the combined rate of incidence of nephrotoxicity overall regardless of cisplatin frequency in our entire population (cohort A and B combined) was 11% which shows a trend toward reduction of nephrotoxicity as historically reported incidence of nephrotoxicity with cisplatin approaches 28% ^1,2^. Unfortunately, historical rate is based on data from multiple cancers and cisplatin dosing regimens so it is cannot be directly statistically compared to our patient population. This apparent improvement in nephroprotection regardless of cisplatin frequency against historical controls could be due to the use of metformin in 25% of our patient, or it could be attributed to the use of a combination of hydration, mannitol, and magnesium sulfate infusions for nephroprotection during cisplatin treatment. In general, hydration is always indicated for nephroprotection in patients treated with platin compounds^34–36^. Second, mannitol infers renal protection against cisplatin toxicity by increasing cisplatin elimination rate while decreasing its concentration in urine through osmotic diuresis^37,38^, but it can also contribute to increased nephrotoxicity in patients with hypertension and diabetes^37^. However, this increase in rate was not seen in cohort A, but it might have reduced our ability to detect a statistically significant difference between the groups since 88% of patients in cohort A were diabetics. Third, magnesium sulfate was shown to have a nephroprotective effect against cisplatin induced nephrotoxicity which is usually exacerbated with magnesium deficiency ^39–42^. This effect is due in part to magnesium sulfate downregulation of OCT2 transporter and upregulation of MATE1 which confers a renal protective effect by preventing accumulation of cisplatin^39^. Hence, metformin could have an additive nephroprotective when combined with magnesium sulfate. However, evidence-based guidelines on specific hydration regimens are limited, and multiple hydration protocols exist among different health systems^43^. Our study is limited by low sample size, and lack of available matched historical controls. nonetheless, our data provides support to the use of this combination of agents for nephroprotection, which supports Crona et al. finding that a combination of hydration, magnesium sulfate and mannitol is the best practice for cisplatin nephrotoxicity protection^44^.

## Conclusion

Our data reaffirms the higher risk of nephrotoxicity for patients on Q3weeks regimen compared to weekly regimen, however, we were unable to show a statistically significant effect in direct comparison between cohort A and cohort B, due mainly to the low number of patients identified. Hence, larger studies in non-diabetic patients are needed to accurately identify an effect.

## Data Availability

All deidentified data produced in the present study are available upon reasonable request to the author after IRB approval of the request

## Acknowledgment

The authors acknowledge the University of Michigan Medical School Research Data Warehouse and DataDirect for providing data aggregation, management, and distribution services in support of the research reported in this publication.” Kheterpal, S. (2015) RDW/DataDirect: a self-serve tool for data retrieval. Available at the University of Michigan at: https://datadirect.med.umich.edu/

