## APPENDIX for "A single center retrospective study to examine the effect of concomitant metformin treatment on cisplatin induced nephrotoxicity in adult HNSCC patients between 2015-2021"

### Appendix A

Table 1. Demographic and Clinical Data for Study Cohorts

|  | Cohort A (n = 18) | Cohort B (n = 54) |
| --- | --- | --- |
| Mean age, y | 56 | 59 |
| Male No. | 18 | 48 |
| Female No. | 0 | 6 |
| Hypertension | 10 | 24 |
| Cirrhosis | 0 | 1 |
| Smoking Exposure | 15 | 34 |
| Diabetes | 16 | 3 |
